# A Data-Driven Evaluation Approach for Assessing Student Nurse Training Effectiveness in Clinical Practice Using A Fuzzy Mathematics Model

**DOI:** 10.1101/2020.09.28.20203570

**Authors:** Yanmei Liu, Yuwen Chen

## Abstract

The overall performance of student nurses during training and subsequent medical treatment practice has a direct effect on the quality of healthcare they provide in hospitals. The evaluation of student nurses’ overall performance is usually not straightforward, as the evaluation criteria includes many aspects and it’s difficult to develop a generic metric. Fuzzy mathematics provides a mathematical tool for processing data with fuzziness. Using fuzzy mathematics theory enables data-driven evaluation of the overall performance of student nurses after their training program.

## INTRODUCTION

Nursing demands much practice. Student nurses need to learn skills and develop positive attitudes and actions in clinical medical treatment practice. As a result, clinical nursing practice is an important topic in nursing education. The quality of clinical nursing practice has a direct effect on healthcare quality. Nursing education needs to train student nurses with high nursing skills. To ensure and improve the all-around quality of student nurses, it is important to establish a system that evaluates the overall quality of student nurses [1, 2].

Data-driven evaluation has been used in evaluating the performance of various clinical procedures and medical treatments [3-16]. In this research, we use a system-based on the data-driven fuzzy mathematics theory [17-23] to evaluate the overall quality of student nurses in medical treatment practice through a quantitative method. In this approach, we turn qualitative indicators into quantitative indicators based on a variety of clinical-relevant indicators.

## THEORY

Fuzzy mathematics is an emerging field after classical mathematics and mathematical statistics. Fuzzy mathematics expand the application of mathematics from precise phenomena to fuzzy phenomena. It mainly researches on the inaccurate inherent cases of things, which reflects the uncertain classification of things arising from the transition between differences. It is not reasonable to classify the overall quality of student nurses into ‘high’ and ‘low’; on the contrary, through the statistical analysis of qualitative indicators, we may get a scientific evaluation. As the overall quality of student nurses is changing over time, evaluating the overall quality of student nurses by evaluating the membership conditions of those indicators affecting nurses’ quality to the overall quality of student nurses will be effective. During the evaluation, indicators under consideration form the indicator set, and the levels of evaluation form the evaluation set [18].

Building a fuzzy general evaluation model contains the following steps:

**Step 1** Determine the indicator set of the objects:

*U* = {*u*_1_,*u*_2_, … *u*_*n*_} are the quality indicators of objects, namely, the Assessment indicators in the evaluation;

**Step 2** Determine the evaluation set:

*V* ={*v*_1_, *v*_2_,…, *v*_*m*_} are levels given to the objects in the evaluation, such as excellent, good, and bad.

**Step 3** Establish the evaluation matrix:

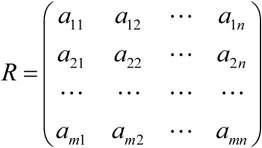

**Step 4** General evaluation:

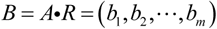

## MATERIALS AND METHODS

### Subjects

208 student nurses were clinically trained in our hospital during 2010. Among those students, 18 are undergraduates. Among them, there are one 23-year-old male and 17 female students. Six of them are 22 years old, eight students are 23 years old and three students are 24 years old. Besides, the participants include 190 junior college students, all females, aging from 18 to 22. Among them, 3 students are 18 years old,18 students are 19 years old, 38 students are 20 years old, 107 students are 21 years old and 24 students are 22 years old. The average age of the students is 20.88±1.05 years old.

### Referees

The referees include 50 nursing teachers and 50 patients, who evaluated the overall quality of student nurses. Among the nursing teachers, one teacher is the deputy director of nurse, 21 teachers are staff nurses, and 28 teachers are ordinary nurses. All of them have work over 8 years. In terms of their teaching experiences, 22 teachers have teaching experience of more than 5 years, 28 teachers have teaching experience of about 3 to 5 years. 12 teachers are leaders of the nurse and 38 teachers are clinical caregivers. Patients involved in our research come into the hospital for over 5 days; all have a degree higher than junior high school. The patients were able to read and understand our questionnaires.

#### Establishing the mathematic model of the overall quality evaluation system of student nurses

According to the general fuzzy evaluation model, we need to generally analyze indicators which affect student nurses’ quality and establish the indicator system with the help of reasonable evaluation indexes. Then we determine the index proportion through multi-factor statistical methods according to evaluation criteria, standardized evaluation indexes, and the qualified membership function. Finally, we combine the index qualified data and proportion vector, getting a total score, with which we then evaluate the overall quality of student nurses.

#### Establishing the evaluation indicators set

The overall quality of student nurses is mainly determined by their basic quality of career and clinical skills. Both of the two dimensions involve in several indicators. We determined the indicator set of the objects *U* = _{_*u*_1_,*u*_2_, … *u*_*n*}_ considering most of the factors affecting the quality of student nurses according to their practice situation, in order to completely evaluate the quality of student nurses. Considering the fact that there are several factors, this research uses a sub-model containing several sub-indicators, as the following graph shows:

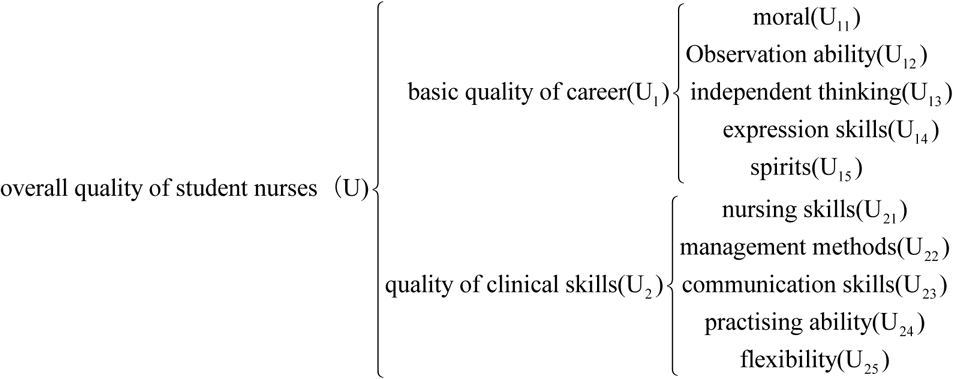

In this graph, the Primary factor set is defined to be

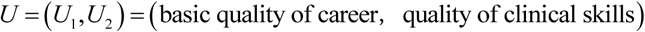

The sub-factor sets are defined to be

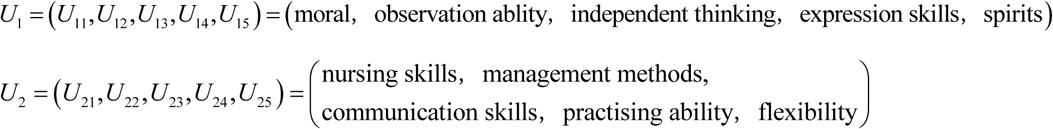

As shown above, there are 2 primary indicators and 5 sub-indicators in each primary indicator.

### Determining the evaluation set

The evaluation set, also called the determination set or the decision-making set, is the evaluation level of every factors in the factor set. Assuming that the evaluation set *V* = {*v*_1_,*v*_2_, …, *v*_*m*_} can be divided into 4 levels in the evaluation model; they are excellent-quality, good-quality, middle-quality and low-quality, as follows:

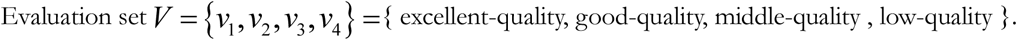

### Single-factor evaluation

We gave out 100 questionnaires to patients and teachers of student nurses in department of cardio-thoracic surgery, department of orthopedics, department of pneumology, department of Cardiology, department of Gastrointestinal surgery. Patients and teachers use the same questionnaires, 50 pieces for each. The teachers filled in the questionnaire on the scene, which were all collected immediately. Besides, questionnaires given to patients were delivered with thorough explanation; only 1 patient would receive the questionnaire per room, which was collected 20mins afterwards; 45pcs are collected. In a sum, 95pcs questionnaire were collected and 85 of them are valid and helpful. We get the single evaluation of the 10 sub-indicators belonging to the two primary indicators after analyzing the 85 valid questionnaires.

Evaluating the five factors in the factor set:

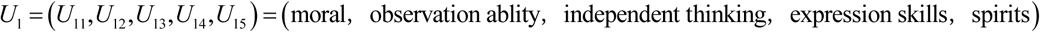

we have:

| $U \backslash V$ | $V_1(\text{excellent})$ | $V_2(\text{good})$ | $V_3(\text{average})$ | $V_4(\text{bad})$ |
| --- | --- | --- | --- | --- |
| $U_{11}(\text{moral})$ | 22 | 51 | 12 | 0 |
| $U_{12}(\text{observation ability})$ | 15 | 44 | 26 | 0 |
| $U_{13}(\text{independent thinking})$ | 11 | 44 | 25 | 5 |
| $U_{14}(\text{expression skills})$ | 22 | 41 | 16 | 6 |
| $U_{15}(\text{spirits})$ | 16 | 41 | 25 | 3 |

This is the single evaluation matrix 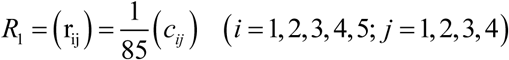 when conducting single-factor evaluation of the five factors in the primary-indicator factor *U*_1_, that is to say, establishing fuzzy maps. *c*_*ij*_ is the ballot number *v* _*j*_ that *u*_*i*_ get. Then we have:

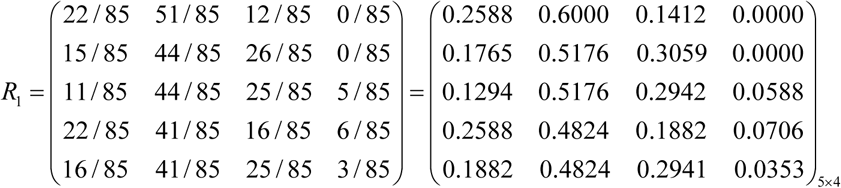

Evaluating the five factors in the factor set:

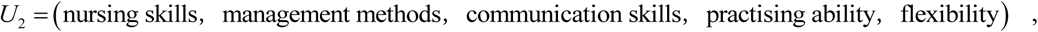

we have:

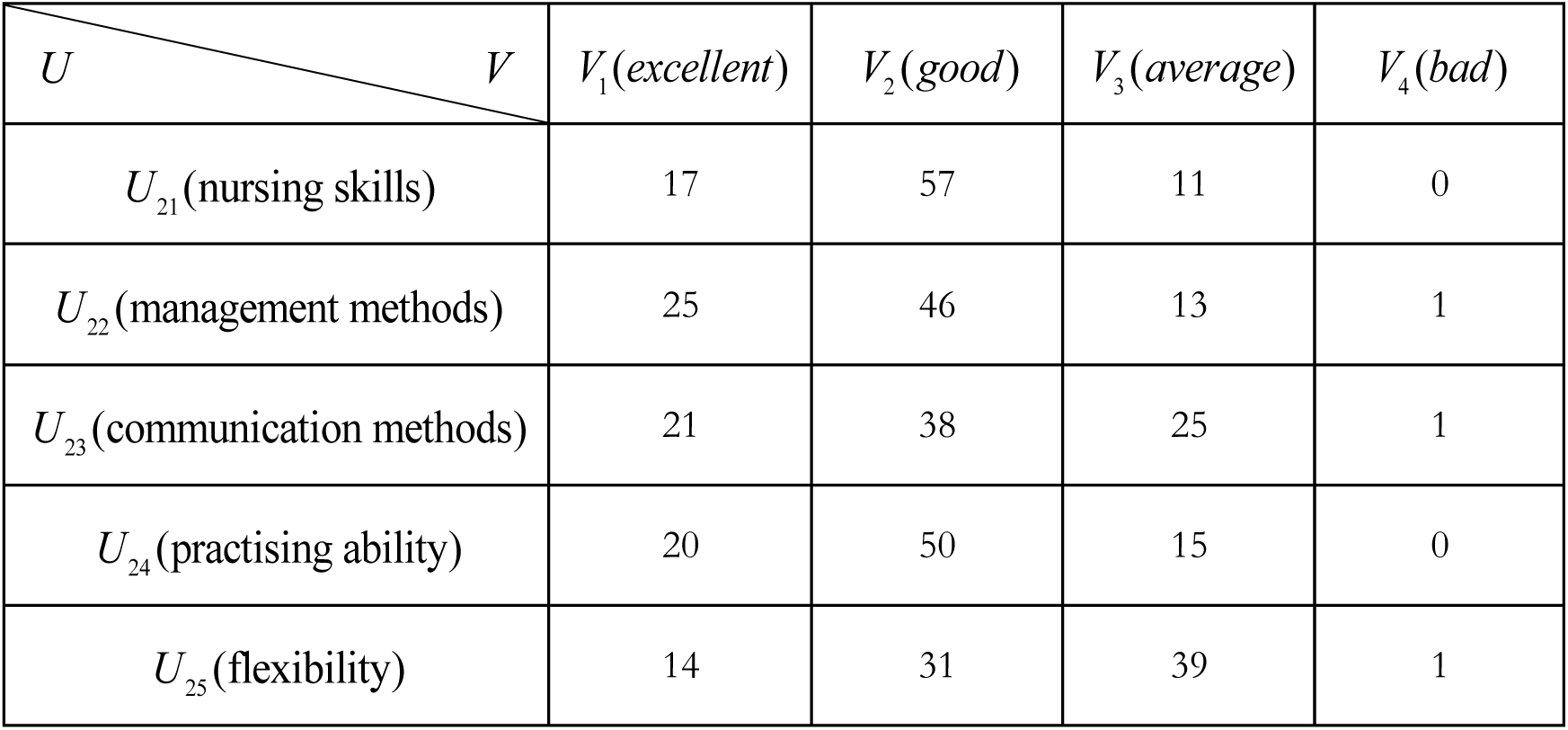

This is the single evaluation matrix 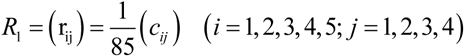 when conducting single-factor evaluation of the five factors in the primary-indicator factor *U*_2_, that is to say, establishing fuzzy maps. *c*_*ij*_ is the ballot number *v* _*j*_ that *u*_*i*_ gets. Then we have:

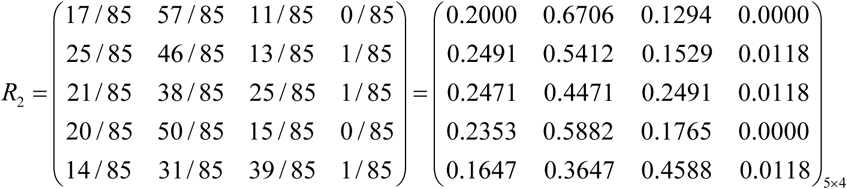

#### Determining the indicator portions

We assign different scale factor to show the importance of different objects in the general fuzzy evaluation procedure. This is called weighting. The scale factor is affected mainly by the importance of the objects. In this passage, different indicators differ from their effects on the evaluation of the overall quality of student nurses. For instance, in terms of the two primary indicators, “clinical skills” is relatively more important; what’s more, among qualities of clinical skills, “nursing skills” is also more important. We need to do research through questionnaires to determine the circumstances above. As a result, by assign the scale factor, the effect of the major factors can be enhanced, which adds to the accuracy of our research.

Assuming that the weighting set of primary indicators is A, we score the two primary indicators through questionnaires, then we determine the weighting according to the score. Because of the two primary indicators, *A* = (*a*_1_, *a*_2_). In the equation, 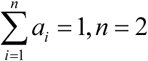 is the weighting set, that is, the fuzzy weighting vector. *ai* is the weighting of the *i*th primary indicator.

In this research, we determine the scale factor through questionnaires. Specifically, we ask the evaluators to score the importance of the three indicators, which makes this research much more scientific. We definite “very important” 4 points, “a little important” 3 points, “so-so” 2 points, “not important” 1 point. Then, we calculate the portion of each score and determine the weighting.

## RESULTS

Through questionnaires, we have:

| choice<br>indicator | Very<br>important | A little<br>important | So-so | Not<br>important |
| --- | --- | --- | --- | --- |
| Basic career quality | 81 | 23 | 1 | 0 |
| Quality of clinical skills | 75 | 8 | 2 | 0 |

According to this, we can work out the score of each primary indicator. They are: Basic career skills 395 points, clinical skills 328 points. As a result, we can determine the weighting of each choice according to the proportion of each primary-indicator score:

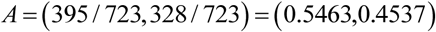

Similarly, we assume that the weighting sets of sub-indicators are *A*_1_, *A*_2_. Then, there are: *A*_1_ = (*A*_11_, *A*_12_, *A*_13_, *A*_14_, *A*_15_), *A*_2_ = (*A*_21_, *A*_22_, *A*_23_, *A*_24_, *A*_25_), in it,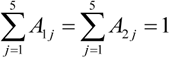.

For the determination of sub-indicators, we get the statistics table with the help of the questionnaires. We set “NO.1” 5 points (which means the most important), set “NO.2” 4 points, “NO.3” 3 points, set “NO.4” 2 points, and set “NO.5” 1 point (which means the least important).

The ranking of the five sub-indicators below “basic career quality” is as follows:

| Ranking<br>Indicator | NO.1 | NO.2 | NO.3 | NO.4 | NO.5 |
| --- | --- | --- | --- | --- | --- |
| Moral | 56 | 10 | 6 | 9 | 4 |
| Observation ability | 14 | 15 | 25 | 24 | 7 |
| Independent thinking | 4 | 15 | 16 | 28 | 22 |
| Expression ability | 1 | 16 | 27 | 12 | 29 |
| Spirits | 9 | 29 | 12 | 12 | 23 |

The scores of the five indicators are 360, 260, 206, 203, 244. According to the proportion of those scores, we can get the weighting:

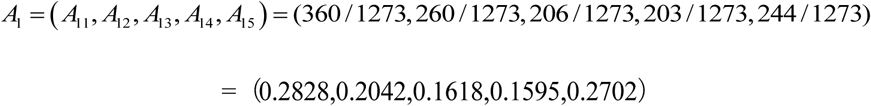

In a word, weighting is the most important part of fuzzy general evaluation, which reflects the status of every factor. In this project, almost any professor can hardly show an accurate data because of the large number of factors involved. Considering this case, we choose questionnaire (multi-factor statistic methods) to determine the weighting, which is more scientific and has little effect on the evaluation result.

### General evaluation of the sub-factor set

In this passage, we use weighted statistical method to estimate the weight of each factor in the sub-factor set. With the former result that the weighting set of the five factors in *U*_1_ is *A*_1_ = (0.2828,0.2042,0.1618,0.1595,0.2702), we generally evaluate the sub-factor set *U*_1_ = (moral,observation ablity,independent thinking,expression skills,spirits), calculate with *M* (·, +), getting 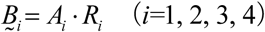.

The general evaluations on the career basic quality are as follows:

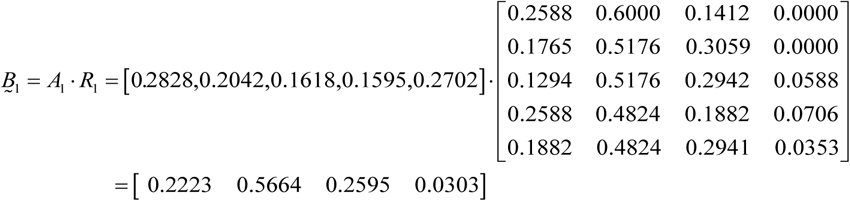

Similarly, evaluate on the sub-factor set

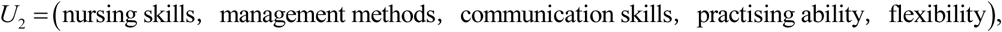

we have:

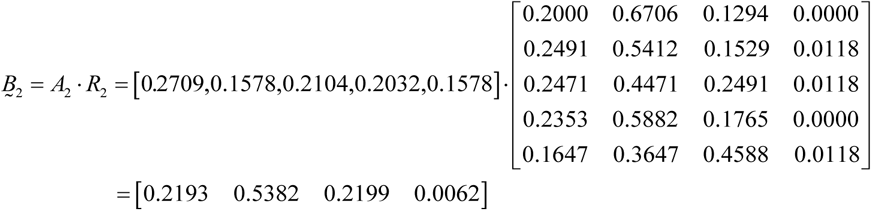

### General evaluation of the primary factor set

After generally evaluate the sub-factor set *U*1,*U*2, we can get the evaluation matrix *R* of the primary set *U* = (basic quality of career,quality of clinical skills), the rows of which are 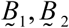, that is:

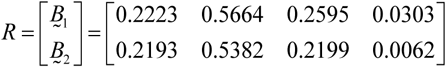

We generally evaluate on it, and have:

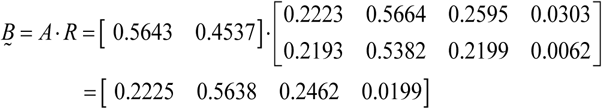

According to the real situation of our survey, we classify the overall quality of student nurses in our hospital into “high”, “good”, “average” and “low”. Then we correspond the four data we get in our overall evaluation with the evaluation set made of these four level *V* =_{_*v*_1_, *v*_2_, *v*_3_, *v*_4}_ ={excellent-quality, good-quality, middle-quality, low-quality}. The final results are 0.5638 > 0.2462 > 0.2225 > 0.0199, so the overall quality of student nurses in our hospital is “good”.

## DISCUSSION

This research shows a novel method in our hospital to evaluate the overall quality of student nurses. We first describe the fuzzy indicators reflecting evaluated things with membership function by constructing a fuzzy membership set, then generally evaluate every indicator using Fuzzy transformation theory. We use a quantitative method, which turn qualitative indicators into quantitative ones and consequently improve the accuracy of our research. This practice provides comparable basis for our hospital to take measures to improve the overall quality of student nurses, which make the decision much more scientific and reasonable. In our research, 2 primary indicators and 10 sub-indicators are used. Through questionnaires, we get the evaluation of the overall quality of student nurses in our hospital from the teachers and patients and then systematically analyze the overall quality of student nurses, which is comprehensive and effective. The final result shows that the overall quality of student nurses in our hospital is “good”, which corresponds to the clinical practice reality and is helpful to the future teaching in our hospital. This method is demonstrated to enable data-processing and analysis of the overall data-driven quality indicator model in the clinical evaluate of the nursing student training quality and effectiveness.

## Data Availability

Data is not available for public access.

